# Samsung Galaxy Watch5 Pro and Withings Body+ generate electrical interference on CRT-Ds

**DOI:** 10.1101/2023.10.05.23296573

**Authors:** Nathan Hansen, Gia-Bao Ha, Penny Colvin, Roger A. Freedman, Antoni Bayés-Genís, Benjamin A. Steinberg, Benjamin Sanchez

**Affiliations:** Department of Electrical and Computer Engineering, University of Utah, Salt Lake City, UT, USA; Department of Medicine, University of Utah Health Sciences Center, Salt Lake City, UT, USA; Department of Cardiology, Heart institute, Hospital Universitari Germans Trias i Pujol, Badalona, Spain; Centro de Investigacion Biomedica en Red Enfermedades Cardiovasculares (CIBERCV), Madrid, Spain; Autonomous University of Barcelona, Barcelona, Spain

## Abstract

Smart consumer devices with bioimpedance sensing technology apply an electrical current to the body for health and wellness. However, whether these smart devices interfere with cardiac implantable electronic devices (CIEDs) remains unknown. We report electrical interference with benchtop testing from the Galaxy Watch5 Pro and smart scale Body+ to cardiac resynchronization therapy device generators from different manufacturers. These results highlight the need of establishing standard testing procedures to assess the safety of smart devices with bioimpedance sensing in patients with CIEDs.

---

We have recently published a study of the possible effects caused by electrical interference from wearable and smart devices using bioimpedance technology in cardiac implantable electronic devices (CIEDs).^1^ Specifically, we performed an initial evaluation to study the level of interference posed by bioimpedance sensing in certain smart watches, smart scales, and smart rings on pacemakers and implantable cardiac defibrillators (ICDs). Our study included computer simulations and benchtop testing using a bioimpedance hardware instrument that allowed us to report the effect of varying frequency, amplitude, and measurement duration during bioimpedance sensing. Due to the lack of guidelines for benchtop safety testing of bioimpedance devices, we based our initial benchtop study on the ISO 14117 standard, the latter used by CIED manufacturers to ensure that their devices are robust to general electromagnetic interference. Our findings revealed a potential risk of these smart devices to disrupt pacemakers and ICDs at certain frequencies and for specific signal amplitudes which had not been previously reported, with levels of induced voltage at the leads that could potentially interfere with the function of CIEDs. However, whether currently-available consumer devices interfere with CIEDs remains unknown. Here, we are the first to report such results through our follow-up benchtop study specifically evaluating two consumer devices: Galaxy Watch5 Pro (Samsung, Suwon-si, South Korea) and smart scale Body+ (Withings, Issy-les-Moulineaux, France).

We first investigated the frequency, amplitude, and duration of the electrical current applied by both devices; this information is not made publicly available by the manufacturers. We connected the driving electrodes of both devices to a reference resistor of 500 Ohms and an oscilloscope. In both devices, the measurement signal was determined to be a sine wave at 50 kilohertz (kHz). The scale and the smart watch applied a current amplitude of 0.7 and 0.35 milliamperes root-mean-square with a total measurement time of 0.7 and 20 seconds, respectively. Next, using a new tissue-interface circuit, we connected the smart devices tested here to the same cardiac resynchronization therapy device (CRT-D) generators and leads we had tested previously (Figure 1 A). This tissue-interface circuit was specifically designed to represent upper- and whole-body impedance values in the sensing frequency range of these consumer devices.^2^ The effect of electrode-to-skin contact impedance on the correct functioning of the tested CRT-D generators was accounted for in our study with two series resistor-capacitor networks connected in each measurement terminal. We programmed the CRT-D generators with the same configuration settings as in Ha et al.^1^ We tested the CRT-D generators with no skin contact impedance, matched skin contact impedance, and unmatched skin contact impedance between terminals of the smart watch and smart scale to investigate best and worst-case scenarios of electrode contact with the body.

**Figure 1.**
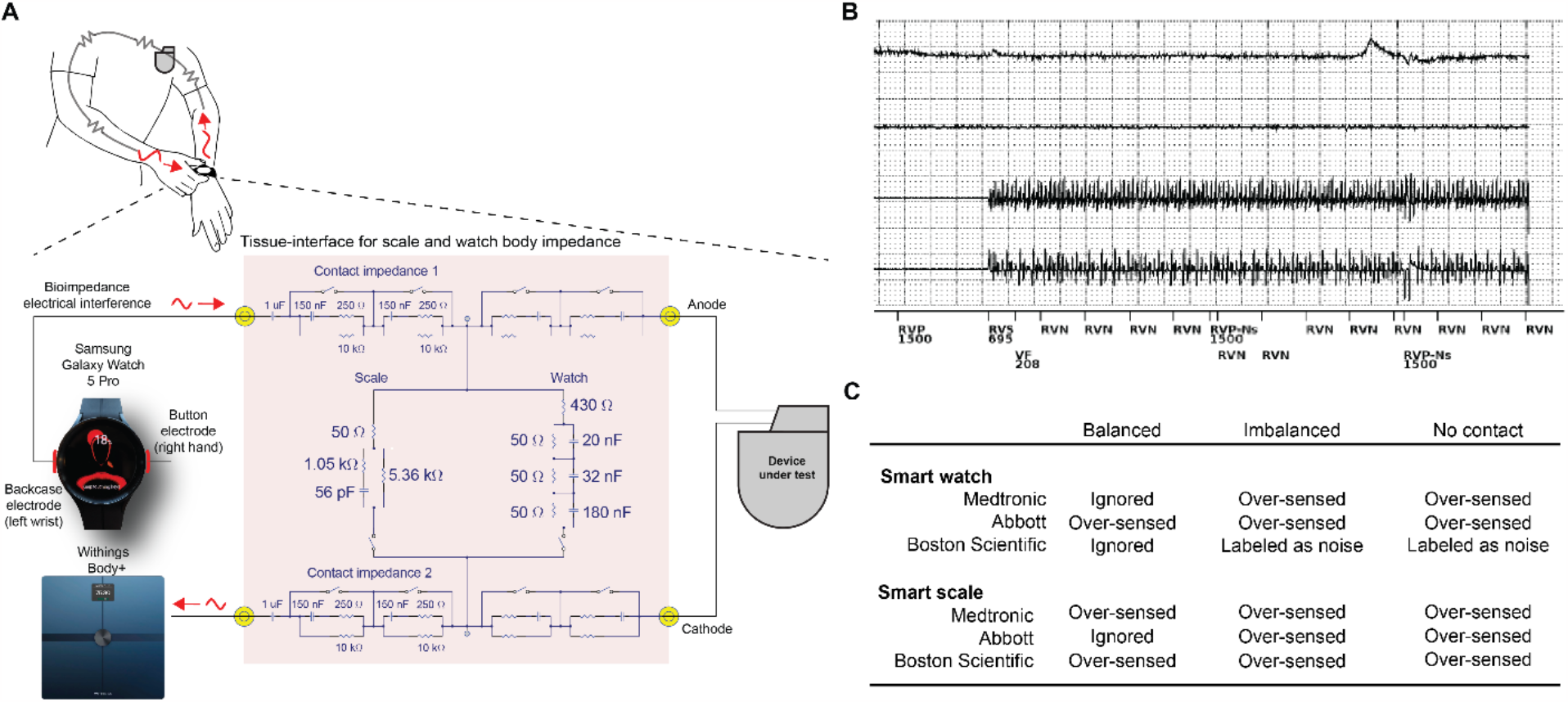
Experimental setup and results for CRT-D response to bioimpedance signals. (A) Schematic showing the testbench setup built to determine the potential of the smart watch Galaxy 5 Pro and smart scale Body Scan to interfere with cardiac implantable electronic devices. The tissue-interface includes electrode-to-skin contact impedances connected to the watch and scale driving terminals. (B) Representative tracing shows appropriate noise detection during smart watch impedance measurement in the imbalanced contact impedance configuration. (C) Summary of interference results considering no contact impedance, matched contact impedance, and unmatched contact impedance.

We observed interference in the form of over-sensing and noise reversion during a smart watch and/or smart scale measurement depending on the vendor and configuration. The smart scale yielded brief signals that were over-sensed by all 3 devices in all 3 configurations, except the Abbott device in a balanced impedance configuration. The smart watch yielded interference more variably: the Abbott device over-sensed smart watch signals consistently; the Medtronic demonstrated over-sensing in all configurations except the balanced impedance (signals were ignored); and the Boston Scientific device appropriately labeled signals as noise except in the balanced configuration (where they were ignored). Sample CRT-D tracing and summary of interference are shown in Figure 1 B and summary of results is shown in Figure 1 C.

This pilot study aimed to evaluate the potential of the commercially available Samsung Galaxy Watch5 Pro and Withings Body+ devices to interfere with CIEDs. Here, we found the bioimpedance sensing functionality of these two smart devices interfered with the correct functioning of CRT-Ds. Our results highlight the imperative necessity of establishing normative testing procedures for scientists, manufacturers, and regulatory bodies to assess the safety of smart devices with bioimpedance sensing in patients with CIEDs. To date, due to the existing regulatory void to evaluate the safety of these devices, manufacturers have opted to use legal disclaimers to prevent patients with CIEDs from using their technologies; yet these patients could benefit from lifestyle and wellness tracking features offered by bioimpedance technology such as monitoring body composition and fluids. Ongoing work is currently focused on expanding the results from our study to other smart devices and CIED models, and to contrast our findings with experimental studies to determine importance in the clinical setting.

## Data Availability

NA

## Funding support and author disclosures

Benjamin Sanchez was supported by an Institutional Research Grant, IRG-21-131-01 Grant DOI #: 10.53354/pc.gr.151058 from the American Cancer Society. He acknowledges the direct financial support for the research reported in this publication provided by the Huntsman Cancer Foundation and the Experimental Therapeutics Program at Huntsman Cancer Institute; the National Cancer Institute of the National Institutes of Health under Award Number P30CA042014 and R21CA273984; the National Science Foundation under Award Number 2319920. Benjamin A. Steinberg was supported by the National Heart, Lung and Blood Institute of the National Institutes of Health under Award Number K23HL143156. The content is solely the responsibility of the authors and does not necessarily represent the official views of the National Institutes of Health.

## Ethical approval and informed consent

Not applicable.

## Notes

**Conflict of interest statement:** Dr. Sanchez is Co-Founder and holds equity in Haystack Diagnostics, Inc., the company has an option to license patented bioimpedance technology where the author is named an inventor. He holds equity and serves as Scientific Advisory Board Member of Ioniq Sciences, Inc., B-Secur, Ltd., and Physio AI, Inc. Dr. Sanchez consults for Myolex, Inc. and Impedimed, Inc., both companies have patented bioimpedance technology where the author is named an inventor. He serves as a consultant to Texas Instruments, Inc., Happy Health, Inc., Analog Devices, Inc., and Eko Health, Inc. Dr. Steinberg reports research support from AHA/PCORI, Abbott, Cardiva, Sanofi, and AltaThera; and consulting to Sanofi, InCarda, Milestone, Pfizer, and AltaThera. The other authors have no conflicts to declare. Dr. Freedman serves on the Cardiac Rhythm Management Medical Advisory Board for Abbott.

### Competing Interest Statement

Dr. Sanchez is Co-Founder and holds equity in Haystack Diagnostics, Inc., the company has an option to license patented bioimpedance technology where the author is named an inventor. He holds equity and serves as Scientific Advisory Board Member of Ioniq Sciences, Inc., B-Secur, Ltd., and Physio AI, Inc. Dr. Sanchez consults for Myolex, Inc. and Impedimed, Inc., both companies have patented bioimpedance technology where the author is named an inventor. He serves as a consultant to Texas Instruments, Inc., Happy Health, Inc., Analog Devices, Inc., and Eko Health, Inc. Dr. Steinberg reports research support from AHA/PCORI, Abbott, Cardiva, Sanofi, and AltaThera; and consulting to Sanofi, InCarda, Milestone, Pfizer, and AltaThera. The other authors have no conflicts to declare. Dr. Freedman serves on the Cardiac Rhythm Management Medical Advisory Board for Abbott.

## REFERENCES

1. Ha G-B, Steinberg BA, Freedman R, Bayés-Genís A, Sanchez B. Safety evaluation of smart scales, smart watches, and smart rings with bioimpedance technology shows evidence of potential interference in cardiac implantable electronic devices. Heart Rhythm. 2023;20(4):561–571.

2. Bogónez-Franco P, Nescolarde L, Bragós R, Rosell-Ferrer J. Measurement errors in multifrequency bioelectrical impedance analyzers with and without impedance electrode mismatch. Physiological Measurement. 2009;30(7):573–587.

